# Development and Validation of the School in China REfusal EvaluatioN Scale (SCREEN): A Tool to Assess School Refusal in Chinese Adolescents

**DOI:** 10.1101/2024.05.31.24307805

**Authors:** Mingzhi Xie, Xiaoyuan Bie, Ye Sun, Guibin Lu, Peng Shu, Yingyu Wang, Mingming Liu

## Abstract

The escalating phenomenon of school refusal (SR) in China has emerged as a significant psychological and educational crisis, with recent surveys revealing a disengagement rate exceeding 70% among students in primary and secondary education. This study introduces the novel School in China REfusal EvaluatioN Scale (SCREEN), specifically designed to assess the multifaceted nature of SR in the Chinese educational context, incorporating culturally relevant socio-psychological variables. Through a robust methodological framework, this research develops and validates the SCREEN, involving the generation and validation of an item pool followed by exploratory and confirmatory factor analyses. The reliability of the SCREEN was affirmed with a Cronbach’s alpha of 0.778, and its validity confirmed through satisfactory factor loadings across multiple dimensions of school engagement. The study’s findings highlight the prevalence and complexity of SR across various educational stages, emphasizing the necessity for holistic intervention strategies that address educational, familial, and societal contributors to SR. The SCREEN’s effectiveness in identifying specific psychological factors contributing to SR underscores the potential for targeted interventions and educational reforms tailored to the unique needs of Chinese students. This research not only contributes to the academic understanding of SR but also provides a practical tool for early detection and intervention, advocating for an integrated approach involving educators, psychologists, and families to mitigate this educational challenge. Future research should focus on longitudinal studies to evaluate intervention efficacy and further refine assessment tools for broader application.

## Introduction

School refusal (SR) and absenteeism are burgeoning crises within the educational and psychological landscapes, presenting complex challenges that impede the healthy developmental trajectories of adolescents worldwide^1,2^. In the context of China, these issues are particularly acute^3,4^, underscored by recent statistics indicating that the rate of school disengagement among middle and elementary school students has escalated alarmingly to 73.3%^5^. This figure emerges from a comprehensive survey involving more than 30,000 students across 500 schools in over 30 major cities, including economically advanced areas such as Beijing, Shanghai, and Hangzhou where rates approach or exceed 80%. These findings suggest a pervasive and deep-seated problem, exacerbated by the high-pressure, competitive nature of the Chinese educational system, which is renowned globally for its rigorous academic demands.

The implications of SR extend far beyond mere academic underachievement. Research has consistently linked SR to a spectrum of long-term detrimental outcomes^6^, including heightened risks of unemployment, compromised social capabilities, and persistent mental health disorders into adulthood. These associations underscore the urgent need for effective strategies that can preempt and mitigate the adverse impacts of SR.

Screening for school refusal among adolescents is a multifaceted process that necessitates a comprehensive consideration of various factors^7,8^. The ubiquity and severity of disengagement across different educational stages highlight that school refusal is not confined to middle school students but spans across age groups, severe among high school females and present even among university students. The diversity of causes—ranging from academic pressure, interpersonal issues, family dynamics, to broader societal factors—underscores the complexity of this phenomenon. Overwhelming academic burdens, insufficient sleep, exam anxiety, dysfunctional peer relationships, and excessively high parental expectations are prominent contributors to students’ aversive attitudes toward school.

Given the foundational role of psychological well-being in educational engagement, it is imperative for educators to actively delve into the psychological realms of adolescents, adopting a humanistic approach to psychological counseling. The application of psychological therapeutic methods, such as integrating innovative content and goal systems in psychological guidance, is crucial in reforming educational modes and methods of psychological support. Additionally, enhancing social support and establishing effective emotional and psychological counseling models are vital for alleviating the disengagement emotions among youths. Implementation of intervention measures, the development of robust study habits, mindful parental management, enhanced classroom management by teachers, and increased regulatory measures on entertainment venues by government entities are all critical in mitigating school refusal.

Further research and the development of more refined tools are necessary due to the current inconsistencies in the conceptualization of school refusal and the lack of reliable instruments for measuring its psychological structure. This indicates an ongoing need for further research and tool development to more accurately identify and address school refusal. In light of these concerns, this study aims to delve deeper into the prevalence and primary drivers of SR among Chinese adolescents. A critical component of our approach involves the introduction of **the School in China REfusal EvaluatioN Scale (SCREEN)**. This innovative diagnostic tool is tailored specifically to the Chinese context, incorporating unique socio-cultural variables that influence educational engagement in this setting. By enhancing the precision and effectiveness of early SR detection, the SCREEN is poised to facilitate timely and culturally congruent interventions. Ultimately, this research seeks not only to elucidate the factors contributing to SR in China but also to pioneer effective tools and methodologies that can be adapted for global use in combating this critical educational and psychological issue.

## Methods

### The School in China REfusal EvaluatioN Scale (SCREEN) scale construction

This study was designed to identify and organize the indicators of SR, and to develop the novel scale. A two-stage approach was followed: (1) generating an item pool, examining the content validity of these items using experts reviewing, and then scale construction, and (2) identify relevant manifestations of school refusal using the content analysis method. The SCREEN aversion to study questionnaire is designed not only to identify symptoms of test anxiety but also to capture a broader spectrum of academic disengagement (supplementary file 1).

The SCREEN aversion to study questionnaire is a comprehensive instrument comprising 50 items designed to evaluate the multifaceted aspects of academic disengagement. Items 21 to 37 are derived from the Sarason Test Anxiety Scale (TAS), which is internationally recognized as one of the preeminent measures of examination-related anxiety. The TAS has been extensively validated and is known for its robust psychometric properties. The remaining items of the SCREEN questionnaire are stratified into three distinct domains: aversion to examinations, functional aversion during lectures as mapped by the TAS (Items 18-20, and Item 44), and aversion to completing homework assignments (Items 45-49 and Items 38-43). These domains encapsulate the breadth of academic avoidance behaviors and attitudes that have been empirically linked to decreased academic performance and well-being.

Furthermore, the SCREEN questionnaire incorporates a set of items that address common concerns frequently articulated in educational counseling and consultation sessions (Items 1-17). These items probe into the students’ intrinsic attitudes towards learning, encompassing their sense of meaning and purpose in their academic pursuits. Such inclusion enriches the SCREEN questionnaire’s capacity to discern underlying motivational factors that may contribute to the academic aversion, thus providing a more nuanced understanding of the students’ educational experience.

### Procedure and participants

Generally, parents of 71 children with SR were recruited through social media groups for parents of children with SR, and the authors’ professional and social networks. The parents were asked to complete an anonymous, online questionnaire, including demographic information and the **SCREEN**. The study was prospectively reviewed and approved by the Centers of MingShiYouXin and MindSage for Research Data. In compliance with ethical guidelines, informed consent was obtained from all participants or their legal guardians before their inclusion in the study. This information was thoroughly documented, adhering to both ethical standards and the requisite legal requirements concerning research involving minors.

### Data analysis

In the development of the SCREEN scale, our data analysis approach was comprehensive, employing a series of statistical techniques to ensure the reliability and validity of the instrument. The initial step involved calculating Cronbach’s alpha coefficients for the overall scale and each distinct subscale to assess internal consistency. We aimed for alpha values of 0.7 or higher, which are indicative of good reliability. This was supplemented by examining the corrected item-total correlations (CITC) for each item, ensuring that each contributed positively to the coherence of the scale.

For further validity analysis, we conducted an exploratory factor analysis (EFA) to investigate the underlying factor structure of the SCREEN scale. We utilized the Kaiser-Meyer-Olkin (KMO) measure and Bartlett’s test of sphericity to evaluate the data’s appropriateness for this analysis. This phase helped in establishing construct validity by revealing the dimensions within the scale that correspond to different aspects of school refusal. We further assessed convergent and discriminant validity through correlations with established measures of similar and distinct constructs, respectively, to confirm that the SCREEN accurately measures school refusal while distinguishing it from other psychological constructs.

## Results

As shown in Table 1, it can be observed that the reliability coefficient is 0.778, which exceeds the threshold of 0.7, thereby indicating that the reliability quality of the research data is good. With regard to the “alpha if item deleted,” the deletion of any item does not result in a significant increase in the reliability coefficient, suggesting that there is no need to remove any item. Concerning the “CITC values,” the item 37, which corresponds to a dislike for courses where teachers prefer ‘surprise attack’ exams, has a CITC value below 0.4. If this is a pilot test analysis, this item could be revised before collecting formal data (if the analysis is performed on final data, the item could be either deleted or retained). In summary, with the reliability coefficient value exceeding 0.7 and considering the aforementioned factors, the data reliability quality is high and suitable for further analysis.

**Table 1.** Reliability analysis and Cronbach alpha test.

| Items | CITC | Alpha if item deleted | Cronbach's alpha |
| --- | --- | --- | --- |
| Item 5 I believe studying is a grueling task. | 0.679 | 0.719 | 0.778 |
| Item 12 Even if I have nothing to do, I am unwilling to study. | 0.625 | 0.729 |  |
| Item 17 I am very interested in computer games, pad games, etc. | 0.409 | 0.765 |  |
| Item 32 Reviewing for important exams seems to be a major challenge for me. | 0.430 | 0.763 |  |
| Item 37 I dislike courses where teachers like to conduct "surprise" exams. | 0.350 | 0.775 |  |
| Item 41 During homework, I find myself thinking about things unrelated to the homework content. | 0.489 | 0.753 |  |
| Item 44 Reviewing important class content seems to be a major challenge for me. | 0.458 | 0.758 |  |
| Item 47 I dislike teachers who assign homework on content not covered in class. | 0.412 | 0.766 |  |

Standardized Cronbach's alpha coefficient: 0.778. CITC, corrected item-total correlation.

Validity studies are employed to analyze whether research items are reasonable and meaningful. Such studies utilize factor analysis as a method of data examination, conducting comprehensive assessments through various metrics such as the Kaiser-Meyer-Olkin (KMO) measure, communalities, the percentage of variance explained, and factor loadings. These indicators collectively facilitate the verification of the data’s validity level. In the current study, the KMO measure is utilized to determine the appropriateness of extracting information, where a higher KMO value indicates the adequacy of the sample for conducting the factor analysis. Communalities are leveraged to exclude irrational research items, and the percentage of variance explained elucidates the level of information extraction. Factor loadings gauge the relationship between factors (dimensions) and the corresponding items.

As indicated in the **Table 2**, all research items have communalities greater than 0.4, suggesting that their information can be effectively extracted. Additionally, the KMO value is 0.724, which exceeds the acceptable threshold of 0.6, indicating that the information extraction from the data is efficacious. Furthermore, the individual factors explain 27.135%, 22.448%, and 20.957% of the variance, respectively, and the cumulative variance explained post-rotation is 70.540%, surpassing the 50% benchmark. This signifies that a substantial amount of the information content within the research items can be effectively captured.

**Table 2.**
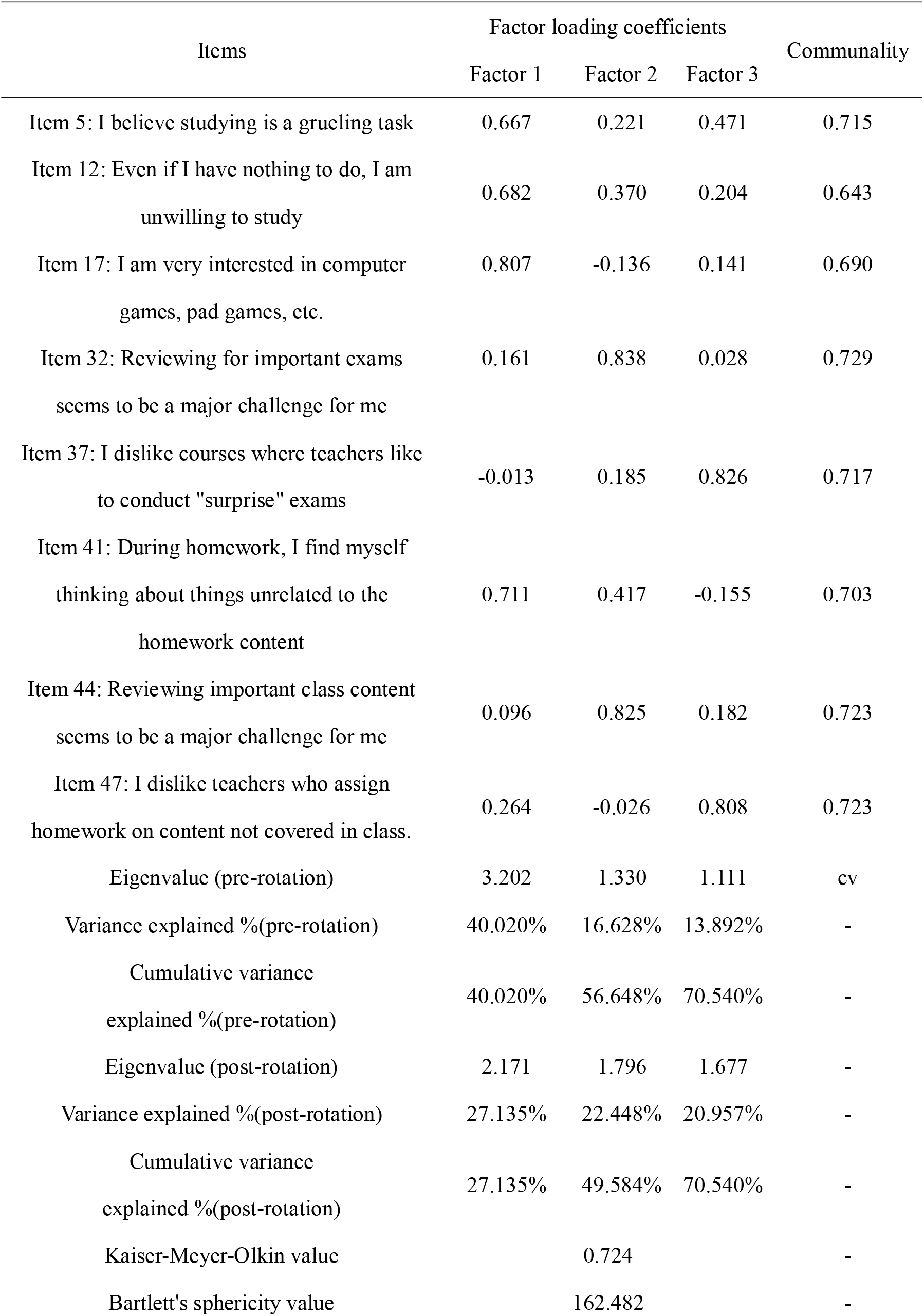
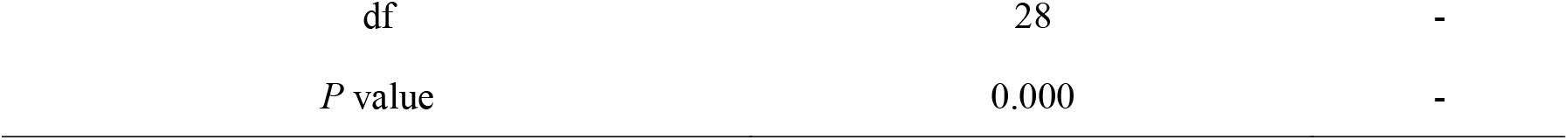
Factor loading of SCREEN items (A final three-factor solution).

Additionally, it is essential to corroborate the correspondence between the factors (dimensions) and research items using the factor loadings. If the relationships align with expectations, the construct exhibits validity; otherwise, adjustments must be made. Factor loadings with absolute values greater than 0.4 are indicative of a meaningful relationship between the items and their associated factors.

The exploratory factor analysis (EFA) conducted on the SCREEN scale yielded substantial insights into its underlying factor structure, affirming the scale’s construct validity. The Kaiser-Meyer-Olkin (KMO) measure was 0.724, indicating a satisfactory level of sampling adequacy for the analysis. Additionally, Bartlett’s test of sphericity confirmed the presence of significant correlations among the items, justifying the suitability of factor analysis with a significance level well below 0.05. The EFA revealed a three-factor solution that accounted for a cumulative variance of 70.540%, with individual factors explaining 27.135%, 22.448%, and 20.957% of the variance, respectively. This robust factor extraction suggested the multidimensional nature of the SCREEN scale. The factor loadings indicated strong associations of specific items with respective factors, exemplifying clear and interpretable factor structures. For instance, Item 5 (“I believe studying is a grueling task”) demonstrated significant loadings on the first and third factors, suggesting its relevance to these dimensions of school refusal. Conversely, Item 37 (“I dislike courses where teachers prefer ‘surprise attack’ exams”) showed a high loading of 0.826 on the third factor, aligning it distinctly with that aspect of the construct. These lines of our results establish a solid foundation for the SCREEN scale’s utility in measuring various facets of school refusal.

## Discussion

The findings of this study elucidate the multifactorial nature of SR among Chinese adolescents, a phenomenon that significantly impacts their psychological and educational development. Through the implementation and analysis of the School in China REfusal EvaluatioN Scale (SCREEN), this research has demonstrated the tool’s robustness and sensitivity in capturing the complex psychological constructs underlying SR. The reliability and validity metrics, particularly a Cronbach’s alpha of 0.778 and satisfactory factor loadings, support the scale’s efficacy in a culturally specific context, thus providing a solid foundation for diagnosing and understanding SR in the Chinese educational environment.

The complexity of SR demands an integrative approach that transcends traditional disciplinary boundaries. By addressing the intertwined psychological, familial, educational, and societal factors, we develop more effective intervention strategies that not only alleviate SR but also promote holistic student well-being and academic success. The data presented in our research unequivocally demonstrate that SR is a pervasive issue that transcends educational stages, affecting a diverse demographic spectrum. This ubiquity of SR necessitates a comprehensive understanding of its multifaceted nature, which involves a confluence of psychological, familial, educational, and societal determinants.

From a psychological perspective, individual factors such as anxiety^9^, motivation^10^, and self-efficacy play critical roles in the manifestation of SR. However, isolating these variables without considering the broader context may lead to incomplete or ineffective interventions. Therefore, it is imperative to adopt a biopsychosocial model that integrates these individual psychological components with external influences. Familial factors, including parental attitudes towards education, socio-economic status, and family dynamics, significantly impact student behavior and attitudes towards learning. Research indicates that supportive and involved parenting can mitigate SR^11,12^, while adverse family environments may exacerbate it. Thus, interventions should consider family-based approaches that promote positive educational environments at home. In the educational sphere, the ethos and practices within schools are pivotal. Teacher-student relationships, curriculum relevance, and institutional support systems are critical in either alleviating or aggravating SR. Educational policies and practices should^13,14^, therefore, be scrutinized and reformed to foster inclusive and engaging learning environments. This may involve teacher training programs that emphasize emotional intelligence and pedagogical strategies tailored to diverse learner needs. Societal factors^15^, including cultural norms, economic pressures^16^, and technological influences^17,18^, also play a significant role. The broader societal context in which students operate can shape their attitudes and behaviors towards education. For instance, socio-economic disparities^19,20^ and cultural attitudes^21,22^ towards education can influence the prevalence of SR. Addressing these societal issues requires cross-sectoral collaboration and policy reforms that aim at reducing inequities and promoting educational equity.

This study’s insights into the psychological dimensions of SR—ranging from academic pressures to interpersonal and familial dynamics—suggest that intervention cannot be monolithic. Instead, a tailored approach that considers the unique circumstances and needs of each student is crucial. For instance, the high correlation between SR and negative perceptions of surprise assessments and unrelatable homework suggests the need for educational reforms^23^, that consider students’ psychological receptiveness and learning preferences. Moreover, the analysis of factor loadings in our study reveals significant insights into how different elements associated with school engagement are perceived by students. For example, items related to direct academic pressures, such as the fear of surprise exams and the challenge of revising for important tests, loaded strongly on distinct factors, suggesting that these areas are particularly salient in the minds of students who refuse school. This specificity indicates potential focal points for interventions, such as modifying exam formats and providing targeted support for exam preparation.

The role of psychotherapeutic interventions, particularly cognitive-behavioral approaches^24^, is reaffirmed by the findings. These techniques, which focus on altering distorted cognition and maladaptive behaviors, are paramount in fostering a more positive school engagement. Given the complex interplay of factors leading to SR, interventions must also extend beyond the individual to include family and school systems. The high incidence of problematic familial interactions related to SR cases highlights the importance of involving families in the therapeutic process, ensuring that family dynamics support rather than hinder educational engagement. Furthermore, the integration of educational and psychological interventions, as suggested by the efficacy of the SCREEN tool, points towards the need for collaborative approaches involving educators, psychologists, and families. Such collaborations could enhance the support system around students, making interventions more comprehensive and contextually sensitive.

In conclusion, this study contributes to a deeper understanding of SR in the Chinese educational context and highlights the importance of a multifaceted approach to address this complex issue. Future research should focus on longitudinal studies to track the efficacy of specific interventions over time and expand the empirical base for developing tailored, culturally sensitive educational and psychological interventions. By continuing to refine the tools and methods used to assess and address SR, stakeholders can better support the educational trajectories and psychological health of adolescents facing this challenge.

## Limitations

The current SCREEN scale is limited by its reliance on self-reported data, which may introduce biases such as social desirability and recall errors, potentially skewing results. Its cross-sectional nature inhibits causal inference, suggesting the need for longitudinal studies to better understand the dynamics of school refusal. The focus on urban, economically advanced areas may not capture the experiences of students in rural or less affluent regions, suggesting a need for the SCREEN’s regional adaptation. Additionally, the study’s sampling strategy does not sufficiently account for the diversity across educational levels and developmental stages of adolescents. Careful attention is required in the translation and cultural adaptation of measurement items to maintain psychological construct validity across cultural contexts. Addressing these issues in future research will enhance the SCREEN’s applicability and contribute to more effective interventions for school refusal.

## Data Availability

All data produced in the present study are available upon reasonable request to the authors.

## Author Contributions

MZX and MML contributed to conception and design of the study. MZX, XYB, YS, and GBL were in charge of data collection. MZX, XYB, YS, and PS planned the statistical analyses. MZX, YS, YYW, and PS performed the statistical analysis. MZX, YYW, and MML wrote the first draft of the manuscript. XYB, YS, GBL, and PS wrote sections of the manuscript. All authors contributed to manuscript revision, read, and approved the submitted version.

## Supplementary Information

### Supplementary File 1. The School in China REfusal EvaluatioN Scale (SCREEN) Test

#### The School in China REfusal EvaluatioN Scale (SCREEN)

consists of 50 items designed to assess students’ emotional responses and attitudes toward school and related academic activities. Each item requires respondents to indicate their level of agreement from ‘Strongly Disagree’ to ‘Strongly Agree.’ The scale explores various dimensions of school refusal, including disinterest in learning, academic stress, school attendance behaviors, and reactions to test-taking situations. This tool aims to help identify emotional barriers that may contribute to school refusal but is not intended for diagnostic purposes. Individuals experiencing significant educational or emotional challenges are encouraged to consult with a mental health professional or school counselor for appropriate support and intervention.

#### School Refusal Emotion Scale (SCREEN) Test

Item 1: Does this statement align with your situation: I find learning completely uninteresting. [Single choice Item]

Item 2: I study only because I am forced to by the circumstances. [Single choice Item]

Item 3: I feel listless when I study. [Single choice Item]

Item 4: In today’s society, there is no benefit in studying. [Single choice Item]

**Item 5: I believe studying is a grueling task. [Single choice Item]**

Item 6: Going to school is a grueling task for me. [Single choice Item]

Item 7: I study solely for my parents. [Single choice Item]

Item 8: I have little interest in studying. [Single choice Item]

Item 9: I feel down and out as soon as class begins. [Single choice Item]

Item 10: I barely understand the content taught by the teacher during class. [Single choice Item]

Item 11: I often copy my classmates’ homework. [Single choice Item]

**Item 12: Even if I have nothing to do, I am unwilling to study. [Single choice Item]**

Item 13: I believe I am not cut out for academic advancement. [Single choice Item]

Item 14: I go to school with my backpack just to kill time. [Single choice Item]

Item 15: I often arrive late or leave early from school. [Single choice Item]

Item 16: I have a tense relationship with my teachers. [Single choice Item]

**Item 17: I am very interested in computer games, pad games, etc**. [Single choice Item]

Item 18: I am unfocused during class and often daydream. [Single choice Item]

Item 19: I feel a headache coming on as soon as I pick up a book. [Single choice Item]

Item 20: I often do things unrelated to studying during class. [Single choice Item]

Item 21: When a major exam is approaching, I always think that others are much smarter than me. [Single choice Item]

Item 22: If I am about to take a test, I feel very anxious beforehand. [Single choice Item]

Item 23: During major exams, I sweat profusely. [Single choice Item]

Item 24: During exams, I find myself thinking about things unrelated to the exam content. [Single choice Item]

Item 25: I often fear failure during exams. [Single choice Item]

Item 26: After a major exam, I often feel so nervous that my stomach feels uncomfortable. [Single choice Item]

Item 27: Doing well in one exam seems to not increase my confidence for the next one. [Single choice Item]

Item 28: During major exams, I sometimes feel my heart beating very fast. [Single choice Item]

Item 29: I always feel very depressed after an exam. [Single choice Item]

Item 30: Before each final exam, I always feel a sense of unease and nervousness. [Single choice Item]

Item 31: During exams, I often get so nervous that I forget things I originally knew. [Single choice Item]

**Item 32: Reviewing for important exams seems to be a major challenge for me. [Single choice Item]**

Item 33: The harder I study for a particular exam, the more confused I feel. [Single choice Item]

Item 34: After an exam, I try to stop worrying, but I can’t. [Single choice Item]

Item 35: If exams could be abolished, I think I could learn more. [Single choice Item]

Item 36: Before major exams, I have no appetite. [Single choice Item]

**Item 37: I dislike courses where teachers like to conduct “surprise” exams. [Single choice Item]**

Item 38: When doing homework in class, I always think that others are much smarter than me. [Single choice Item]

Item 39: If I am about to correct a homework assignment, I feel very anxious beforehand. [Single choice Item]

Item 40: When my mother is around, my palms sweat a lot while doing homework. [Single choice Item]

**Item 41: During homework, I find myself thinking about things unrelated to the homework content. [Single choice Item]**

Item 42: I often fear failure during homework. [Single choice Item]

Item 43: I always feel very depressed after the teacher assigns homework. [Single choice Item]

**Item 44: Reviewing important class content seems to be a major challenge for me. [Single choice Item]**

Item 45: The harder I work on a particular assignment, the more confused I feel. [Single choice Item]

Item 46: If I didn’t have to do homework, I think I could learn more. [Single choice Item]

**Item 47: I dislike teachers who assign homework on content not covered in class. [Single choice Item]**

Item 48: I often feel overwhelmed by the amount of homework I have to do. [Single choice Item]

Item 49: I often procrastinate on starting my homework. [Single choice Item]

Item 50: I often feel stressed when I think about all the homework I have to complete. [Single choice Item]

### Answering System

- Strongly Disagree
- Disagree
- Neutral
- Agree
- Strongly Agree

Please answer the questions by choosing one of the options that best describes your feelings or thoughts.

Note: This scale is designed to measure school refusal behaviors and the emotional experiences associated with those behaviors. It is not a diagnostic tool. If you are experiencing significant distress or difficulties with school attendance, please seek help from a mental health professional or school counselor.

